# Are the sterile insect technique and the incompatible insect techniques effective in reducing *Aedes* mosquito populations?

**DOI:** 10.1101/2024.02.03.24302193

**Authors:** Marie-Marie Olive, Gilbert Le Goff, Thierry Baldet, David Roiz

## Abstract

**Background:** The control of *Aedes aegypti* and *Aedes albopictus* mosquitoes, the main vectors of dengue, chikungunya and Zika viruses, presents several challenges. The difficulties encountered in acquiring funding, implementing measures, obtaining community participation, acceptability and effectiveness, and the problem of insecticide resistance demonstrate the need to develop and optimise innovative vector control strategies. The sterile insect technique (SIT), the incompatible insect technique (IIT) and a combination of both (SIT-IIT) show promise. Numerous trials are being carried out worldwide to obtain evidence of their effectiveness before implementing them in large-scale, integrated vector-control strategies. The main objective of our study is to build an analytical framework for the identification and standardisation of appropriate entomological indicators that could be used to compare the relative effectiveness of the SIT, IIT and SIT-IIT methods in reducing *Aedes* vector populations.

**Methods:** We reviewed the available scientific literature to compare the characteristics, methodologies, effectiveness indicators and results of various trials with the aim of standardising and comparing the indicators used in the trials, such as reductions in the egg hatch rate and in the adult populations.

**Results:** Seventeen trials, either published in peer-reviewed journals or posted as preprints, were selected. We found wide variation among them in experimental design, field implementation and the methods of calculating the indicators. Although limited by the amount of available published data, our results suggest that a reduction in egg hatching greater than 45% results in up to 60% fewer females, greater than 60% results in over 80% fewer females, and greater than 70% results in over 90% fewer females. Therefore, the quality of implementation, assessed on the basis of egg hatch reduction, is statistically associated with effectiveness, assessed on the basis of the reduction in *Aedes* females.

**Conclusion:** We present results suggesting that, when implemented effectively, the incompatible and sterile insect techniques are substantially effective in reducing *Aedes* mosquito populations. Furthermore, these techniques are species specific, non-insecticidal and environmentally friendly. However, it has yet to be shown that they can be scaled up as cost-effective operational tools for vector control and that they substantially reduce arbovirus transmission.

## Introduction

As recent epidemic events, such as the covid-19 pandemic, have made abundantly clear, the generation, accumulation and communication of scientific evidence are key components of reliable policymaking and citizen engagement in public health [1]. The control of *Aedes*-borne diseases, such as dengue, Zika and chikungunya, caused by viruses transmitted principally by *Aedes aegypti* and *Aedes albopictus* mosquitoes is no exception [2,3]. Over the past 50 years these diseases have emerged or re-emerged worldwide and controlling them presents many challenges. Key challenges are the generation of scientific evidence of the effectiveness of *Aedes* control methods and the difficulty of conducting trials to evaluate vector control strategies, especially those concerning epidemiological outcomes in epidemic and endemic areas [2,4,5]. Dengue is the most common arboviral disease, the number of cases having increased 30-fold over the past four decades [3] to around 390 million infections annually with a massive and increasing economic impact [6]. Multiple dengue serotypes are now circulating in all tropical regions causing seasonal epidemics, hyperendemicity, severe haemorrhagic cases and long-term sequelae every year [7]. There have been major epidemics of chikungunya in East Africa and the Indian Ocean (2005-2006) and the Americas and Oceania (2013-2014), and Zika in the Americas (2015-2016) [8–10], while yellow fever has been re-emerging with recent epidemics in sub-Saharan Africa and Brazil [11,12]. In the absence of vaccines and treatments, except for yellow fever and potentially for dengue [13], the prevention and control of *Aedes*-borne viral diseases continues to rely heavily on controlling mosquito vector populations or interrupting human-vector contact through integrated, sustainable, proactive, synergetic vector control management. Integrated vector control uses a combination of several control methods, such as environmental management, source reduction, insecticide spraying or novel vector control methods (for example, sterile insect techniques, *Wolbachia*-based incompatible insect techniques for population suppression or for population replacement and mass-trapping). Public awareness, community mobilisation and intersectoral collaboration should also be included [2]. Focusing on investment in management and control is justified by the fact that both the *Aedes* species *(Ae. aegypti* and *Ae. albopictus)* involved in transmitting these viruses are considered to be among the most economically costly invasive species [14]. While the cost of prevention is an order of magnitude lower than the costs of medical damage and losses, only a modest portion of the total reported expenditure on control management is invested in this area [6]. It exists scarce reliable evidence that integrated vector control strategies are effective, but the few studies that have been rigorously applied show that they can successfully reduce mosquito densities and the transmission of arboviruses [2,15,16]. The effectiveness of such strategies is potentially hampered by inefficient implementation, insufficient coverage and a lack of human, financial and infrastructural capacity and political willingness [2,17]. Of greater concern are the new challenges to *Aedes* vector control, such as the emergence of insecticide-resistant mosquito populations, the increasing aversion of human populations to insecticide treatments and the impact of the latter on non-target fauna and the environment [17]. The development and testing of innovative, non-insecticidal strategies for *Aedes* control has therefore become an urgent priority.

Among the methods developed, the sterile insect technique (SIT), the incompatible insect technique for population suppression (IIT) and strategies combining the two sterilisation techniques (SIT-IIT) show promise [17,18]. Both techniques involve the mass production and release of male mosquitoes that cannot produce offspring. Sterile males released into the field will compete with wild males to breed with wild females. When females mate with a sterile male they will lay sterile eggs that will never hatch. The ultimate goal of these control strategies targeting mosquito populations is to reduce the circulation of mosquito-borne viruses and contain epidemics by reducing vector density.

Trials evaluating the impact of SIT, IIT and SIT-IIT on populations of *Aedes aegypti* and *Ae. albopictus*, the two major vectors of human arboviruses, are currently being conducted worldwide. IIT trials are also being conducted on *Ae. polynesiensis*, a local vector of dengue, lymphatic filariasis and Ross River virus in the South Pacific [18,19]. SIT is being implemented in Europe (Italy, Spain, Greece, Germany, Albania, Montenegro), the Southwest Indian Ocean Islands [20,21] and the Americas (Brazil, Mexico) [19]. IIT alone is being tested in the United States, Australia and French Polynesia [19]. The combined SIT-IIT approach is mainly being implemented in Asia [19,22]. Over the last two decades, about twenty field trials have been conducted worldwide. There would seem to be, therefore, an urgent need to gather together and summarise the increasing volume of evidence and establish a standardised methodology that would allow the entomological effectiveness of these different trials to be compared.

The objectives of the present study are (1) to conduct a systematic review of the quality of the implementation and the entomological effectiveness of SIT and/or IIT field trials to reduce *Aedes* vector populations; (2) to identify appropriate entomological indicators in order to build a framework for evaluating the effectiveness of the various strategies in reducing *Aedes* vector populations; and (3) to present an up-to-date overview of the results of the trials.

## Materials and Methods

### Literature search

Our study focused solely on research articles retrieved via a search of the PubMed and Web of Science databases, supplemented with a search of opportunistic research studies in preprint repositories. Systematic searches were conducted in November 2020 (Supplementary Figure 1), and a complementary search was carried out from December 2020 to April 2022, to include updated publications and preprints on SIT, IIT and SIT-IIT trials. We used the following search strings: in Pubmed: ((“Aedes”[Title/Abstract]) AND (evaluation[Title/Abstract] OR effectiveness[Title/Abstract] OR effect[Title/Abstract] OR assess*[Title/Abstract] OR effic*[Title/Abstract] OR reduc*[Title/Abstract] OR release*[Title/Abstract] OR field study[Title/Abstract] OR randomized control[Title/Abstract])) AND (sterile insect technique[Title/Abstract] OR Wolbachia[Title/Abstract] OR boost*[Title/Abstract])); for ISI in Web of Science: ((“Aedes”) AND TOPIC: (evaluation OR effectiveness OR effect OR assess* OR effic* OR reduc* OR release* OR field study OR randomized control) AND TOPIC: (sterile insect technique OR Wolbachia OR boost*)). The inclusion and exclusion criteria for selecting studies for the systematic review are reported in Supplementary Table 1. Briefly, we included controlled field trials of SIT, IIT (with the exclusion of IIT for population replacement) or combined SIT-IIT for *Aedes aegypti* or *Aedes albopictus* or *Aedes polynesiensis* that targeted eggs and adults, the indicators being the number of adults or eggs per trap [2].

### Description of the study and data extraction

We analysed the trials using a two-step procedure: (1) extraction of the main characteristics of the trials and (2) standardisation of the reduction in the egg hatch rate and the suppression of adults. In the first step, we extracted descriptive information from the articles: type of publication, country and year of trial, technique used, target species, geographical location, epidemiological context, size of the intervention site, release method, total number of sterile males released, duration of the study, indicators used to measure the implementation and effectiveness of the technique and the study conclusions. Details of each recorded or estimated variable are provided in Supplementary Table 2. The data were extracted from each article by one of the authors of the present study using a template specifying the relevant data fields and were then reviewed by a second author.

In the second step, entomological field indicators were categorised into two groups (Figure 1):

**i) Indicators of the quality of implementation**. These describe the quality of the field implementation, such as the density of sterile males in relation to wild males (sterile-to-wild male ratio), the ability of sterile males to mate with wild females (field mating competitiveness), larval productivity and the number of non-viable eggs produced (reduction in egg hatch; Figure 1). From these we standardised a primary indicator of implementation quality: the reduction in egg hatch (also referred to as the percentage of induced egg sterility) using Abbott’s formula [23].

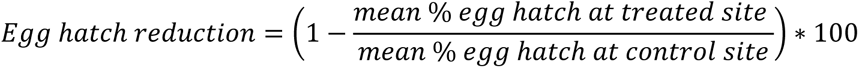

**ii) Indicators of effectiveness**. These describe the ability of the technique to reduce the *Aedes* population, for example, by measuring the density of adults and/or eggs according to different types of trap (Figure 1). From these we standardised one main indicator of trial effectiveness: the suppression of adult *Aedes* females using Abbott’s formula [23].

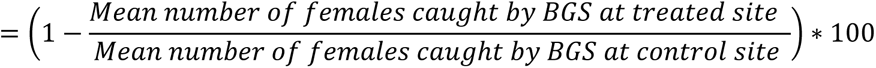

Given that a delay of several weeks is necessary to observe the effect of the method on the mosquito population, we standardised the indicators at three months (month 3) and four months (month 4) after the first release. Data from replicates of the same trial − i.e. those conducted in Australia in 2018 [24], Fresno County, California, USA, in 2018 [25] and Guangzhou, China, in 2016 [22] − were aggregated for the release and control sites. Results from the two control sites in an IIT trial conducted in French Polynesia against *Ae. polynesiensis* were also aggregated. Unfortunately, due to the high heterogeneity of the data, we were unable to standardise the “sterile-to-wild male ratio” indicator (Figure 1).

**Figure 1:**
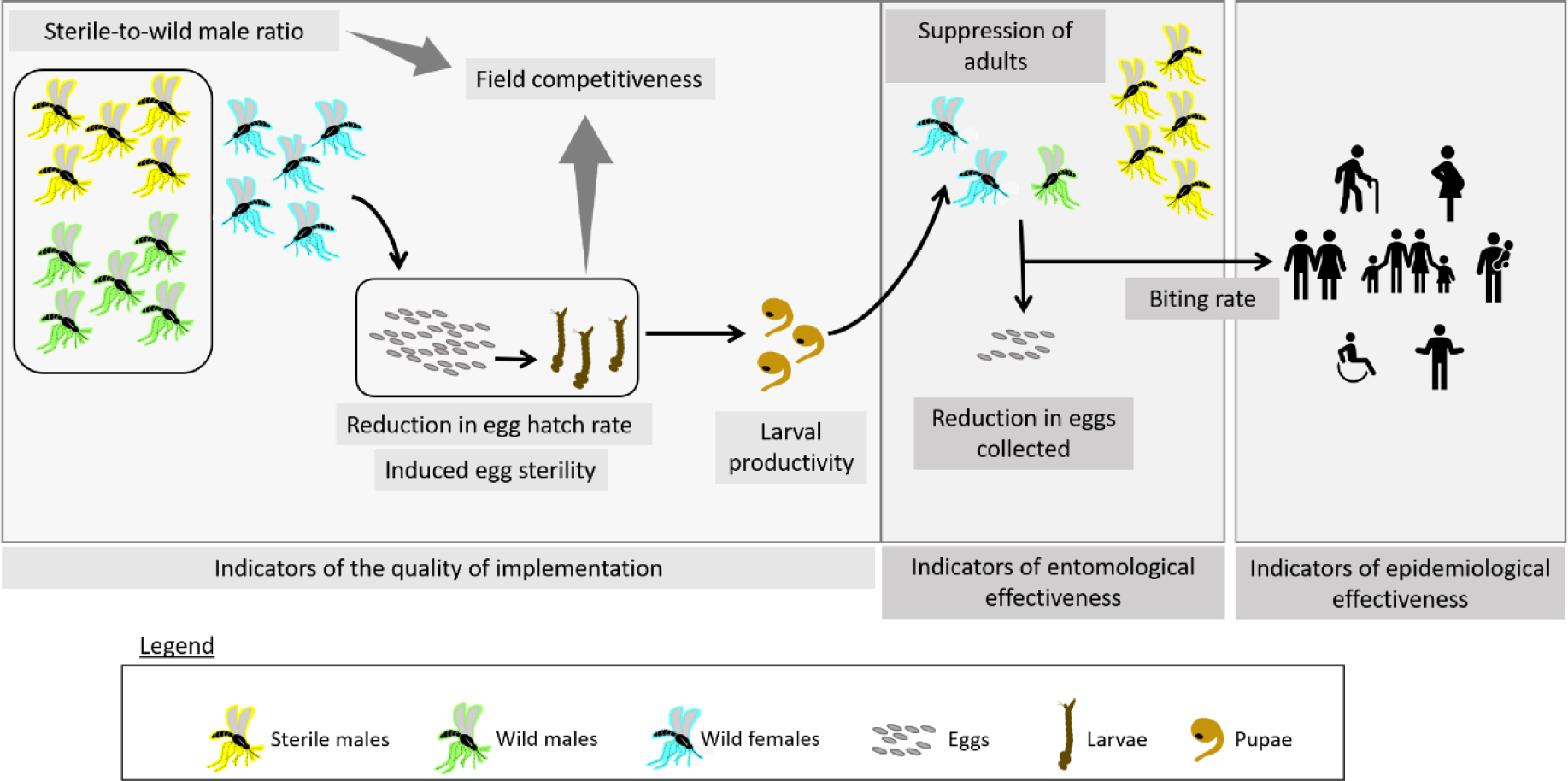
Schematic overview of the indicators used in the trials

Finally, we used a generalised linear model (GLM) and correlation analysis to explore the relationships between the quality of implementation and effectiveness indicators.

The levels of reliability and potential biases of our standardisation were categorised according to three criteria:

- Low, when the original data were not presented at a sufficient level of detail for standardisation. In this case, we extracted data from the figures with the R digitize package [26];
- Moderate, when the data were presented in the manuscript at a sufficient level of detail for standardisation;
- High, when the complete data were presented in the manuscript’s supplementary materials at a sufficient level of detail for standardisation.

To summarise, the following methodology was followed for the study:

1. Title and abstract screening using the PRISMA methodology;
2. Screening of the full text of the papers against the inclusion/exclusion criteria (SI Table 1)
3. Extraction of the main characteristics of the trials including the methods of calculating the quality of implementation and effectiveness indicators (SI Table 2)
4. Extraction of the main results of the trial
5. Selection of the trials with reliable data for standardisation (i.e. with at least monthly data for egg hatch rate and/or female captures)
6. Standardisation of the data at month 3 and month 4

## Results

### Systematic review

Firstly, we identified 828 potentially relevant articles from the searches conducted in November 2020 (Supplementary Figure 1). We ultimately retained 17 studies covering trials of SIT, IIT and SIT-IIT against *Aedes* mosquitoes that had been or were being conducted worldwide. Eight trials focused on *Ae. albopictus*, eight on *Ae. aegypti* and one on *Ae. polynesiensis*. Eight trials involved the SIT technique, six the IIT technique and three the combination of the two techniques, IIT-SIT (SI Table 2).

### Analyses of the trial methodologies

It is of note that we found a range of experimental designs, field implementation strategies and methods for calculating the indicators in the different trials (Table 1). A thorough analysis of the indicators used was therefore necessary to enable us to proceed to an appropriate standardisation.

**Table 1.**
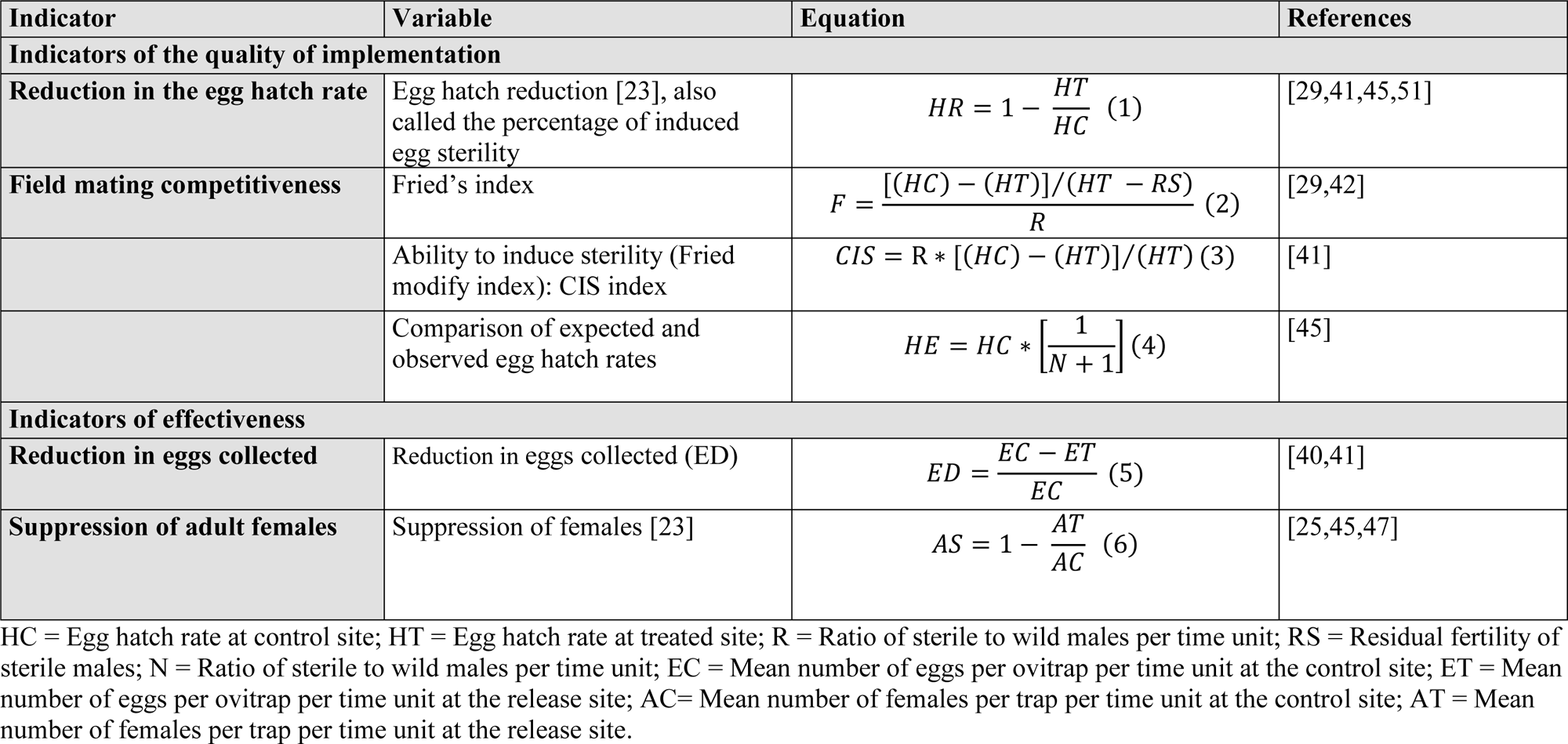
Overview of the indicators of implementation quality and effectiveness in the trials.

First of all, two main indicators of implementation quality were identified: egg hatch reduction and field mating competitiveness of the released sterile male. The egg hatch rate is the most commonly used index of non-viable egg production in the trials. However, this index is calculated by various methods. The first consists in hatching eggs collected in the field in cycles of maturation achieved in the laboratory by drying them then immersing them in a hatching solution [27–29]. With the second method, in addition to assessing the ability of eggs to hatch, unhatched eggs are examined microscopically to see if they are embryonated [21,30]. Egg viability rates are then estimated as the ratio of the number of hatched or unhatched but embryonated eggs to the total number of eggs examined, while the egg hatch rate is calculated as the ratio of the number of hatched eggs to the total number of eggs examined. The third method of measuring egg hatch ability was that proposed by O’Connor et al. [31]: caught females were individualised in oviposition containers and the resulting eggs submerged to hatch; they were then observed for any resulting larvae. The spermathecas were dissected from the females, crushed in a PBS solution on a microscope slide using a coverslip then examined with a compound microscope [31]. In Australia, estimated larval productivity was used rather than egg hatch rate due to the difficulty of assessing viable eggs on sticky ovitraps that were either semi collapsed (potentially viable) or collapsed (nonviable) [24]. The reduction in egg hatch is usually calculated using Abbott’s equation (Equation 1, Table 1).

Field mating competitiveness of sterile males is calculated on the basis of the egg hatch rate, the sterile-to-wild male ratio and the assumption of the residual sterility of sterile males (Equation 2, Table 1). Some authors have suggest omitting residual fertility when it is less than 1% [32] (Equation 3, Table 1) and to calculate instead the ability to induce sterility (Equation 3, Table 1).

The indicators for effectiveness, e.g. the reduction in eggs collected and/or the suppression of females, are usually calculated using Abbott’s formula (Equations 5 and 6, Table 1).

### Data standardisation

Most trials (70%) had sufficient data to normalise either the reduction in the egg hatch rate or the level of female suppression. Both indicators were standardised for seven trials (40%) (SI Table 3). “Egg hatch reduction” was standardised for 10 papers for month 3 and month 4 after the first release (Table 2 and Figure 2). Most “egg hatch reduction” indicators were standardised for SIT (7 for month 3 and month 4 after the first release) and three were standardised for both IIT and SIT-IIT. In contrast, only one indicator of “female suppression” was standardised for SIT, whereas four of these indicators were standardised for IIT and three for SIT-IIT (Table 2 and Figure 2). All standardised indicators for SIT were for *Ae. albopictus,* while the other techniques concerned *Ae. albopictus* or *Ae. aegypti* (Table 2 and Figure 2). The standardised indicators show egg hatch reduction ranging from 17% to 70% and female suppression ranging from 11% to 88% (Table 2 and Figure 2).

**Table 2:**
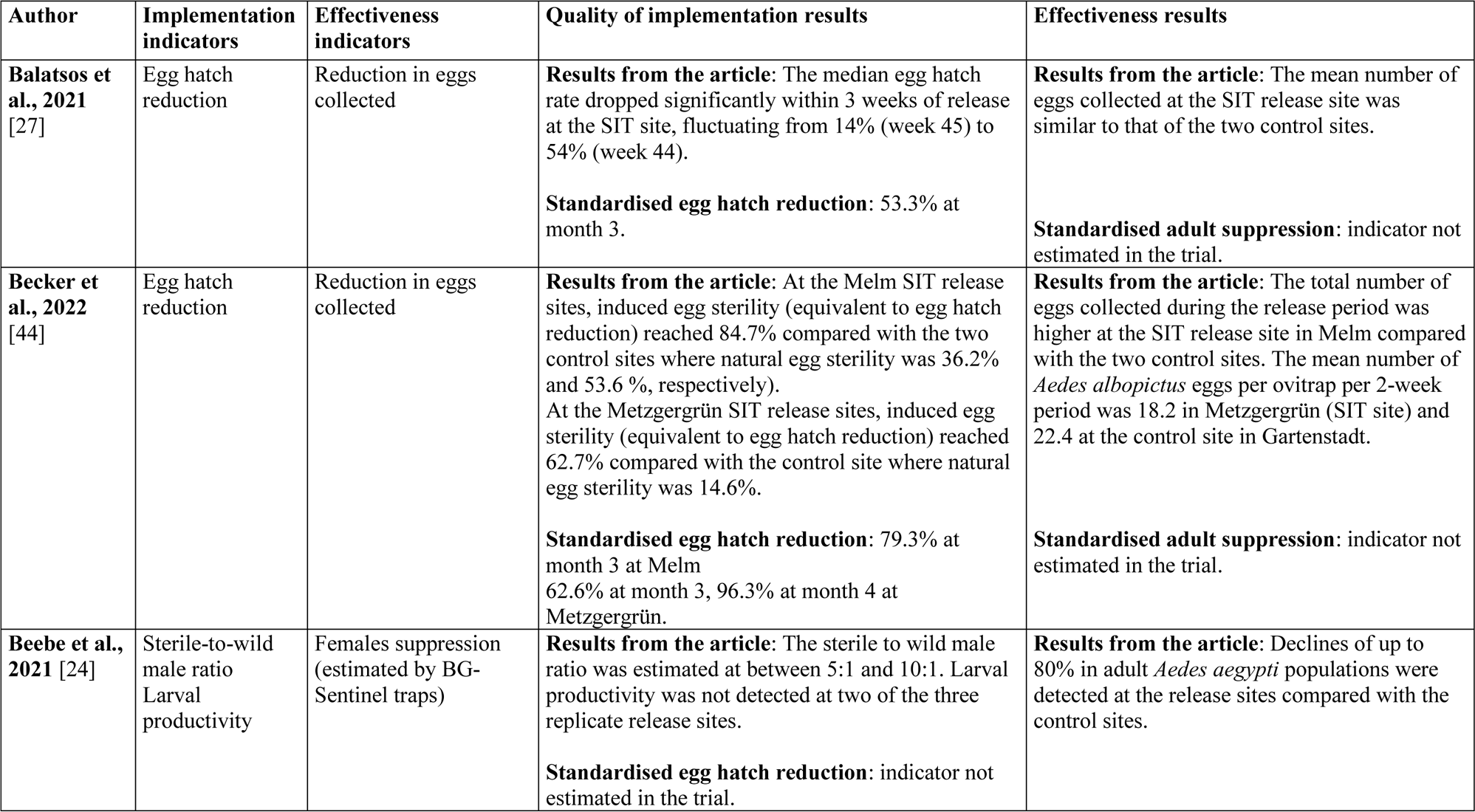
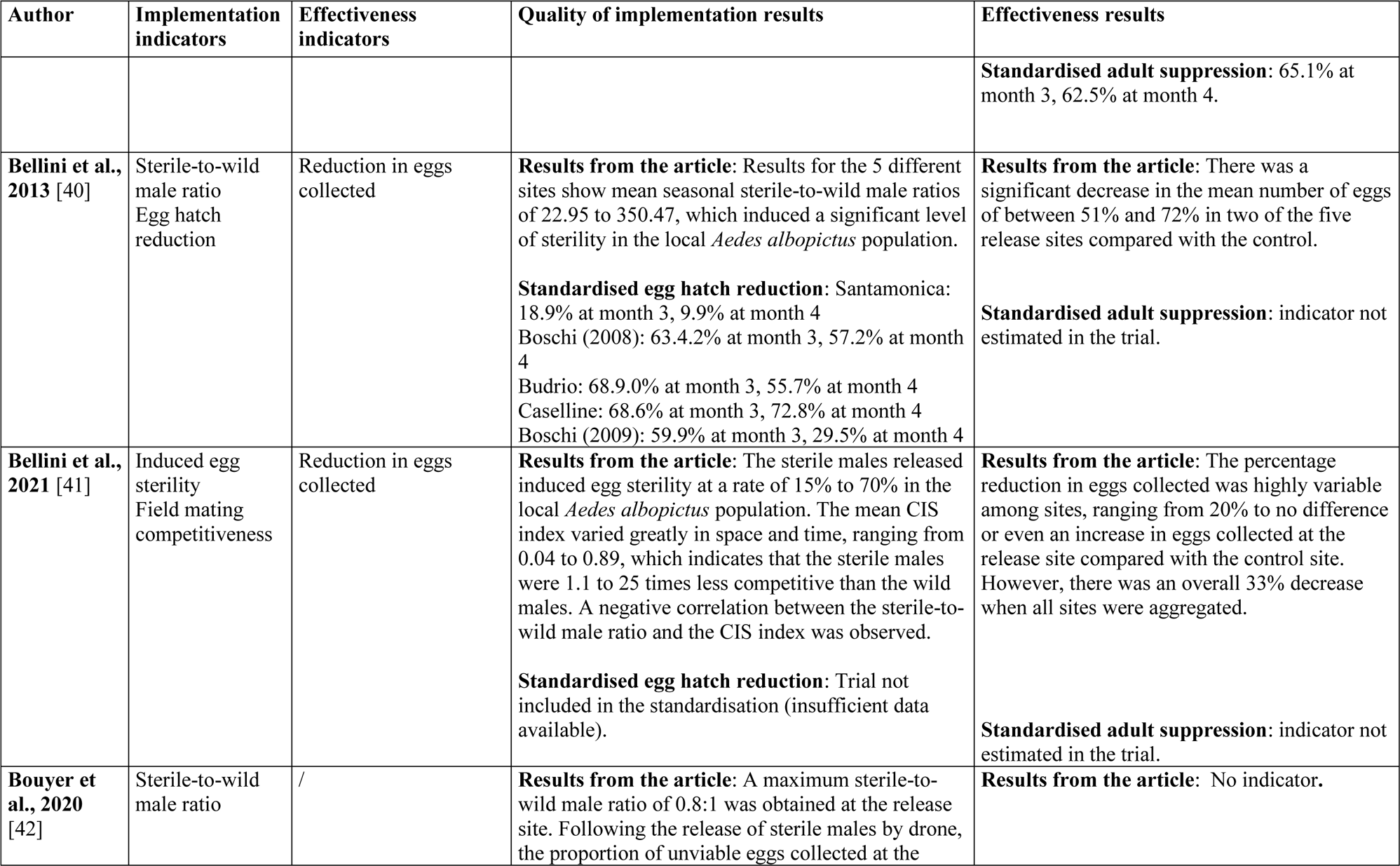
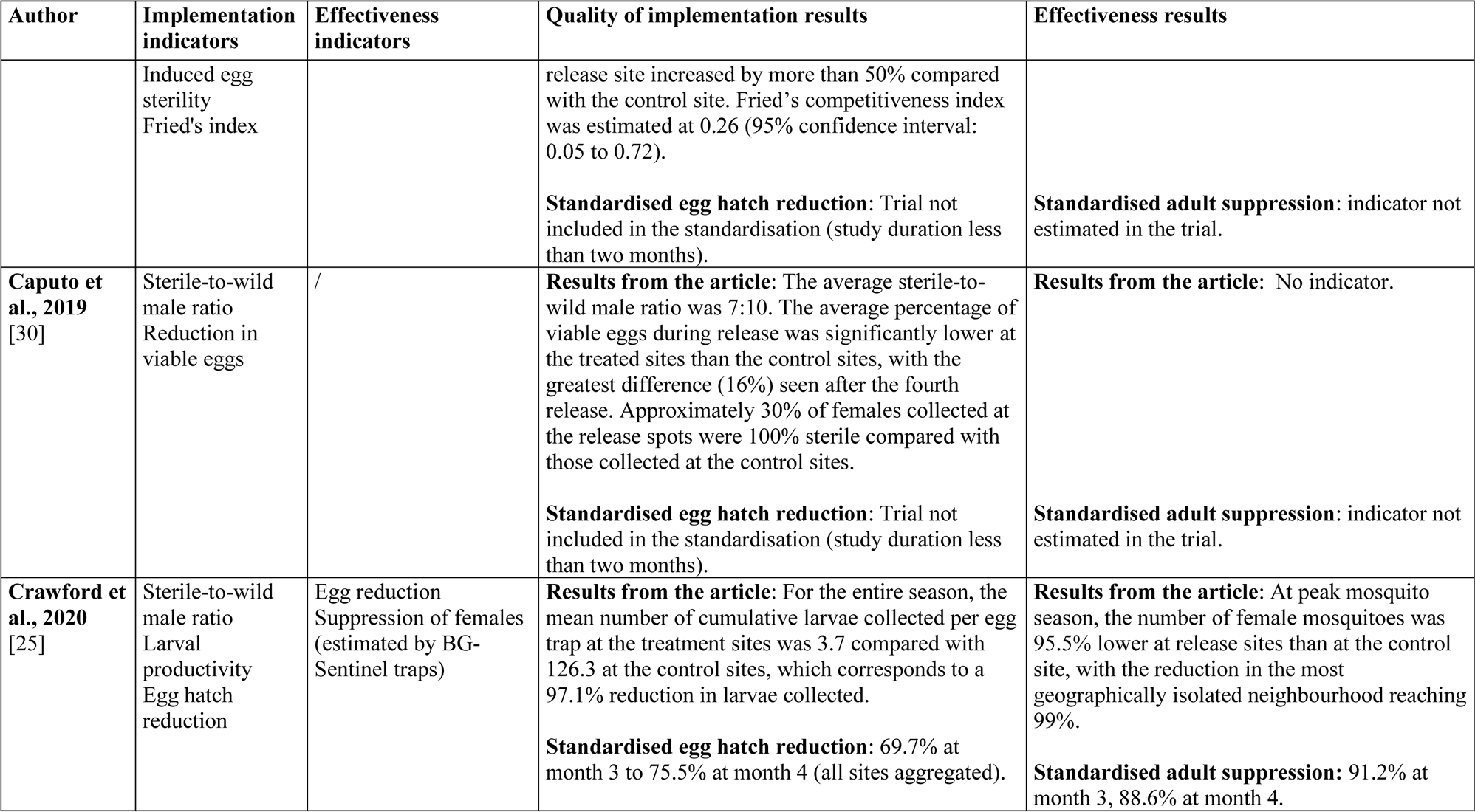
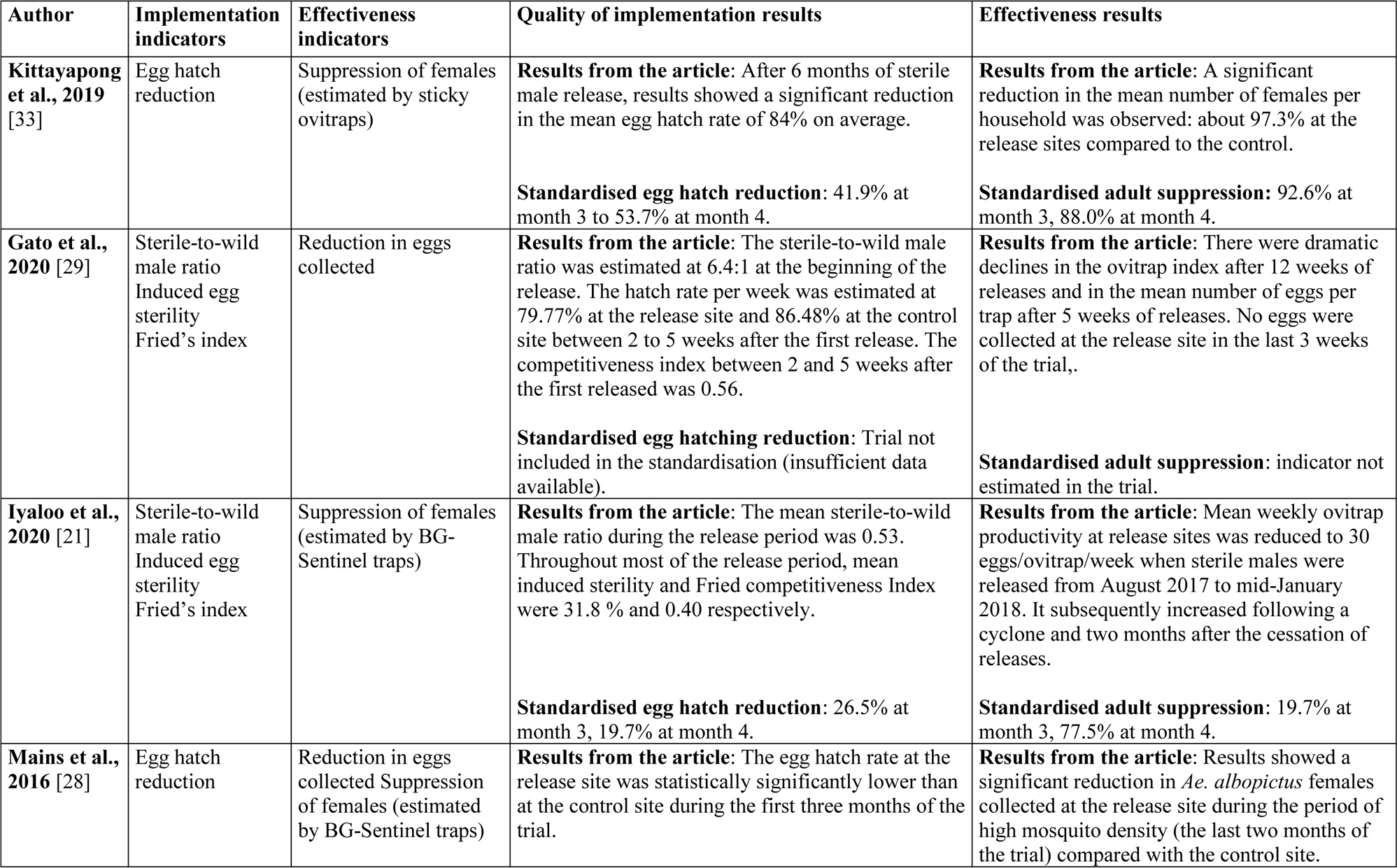
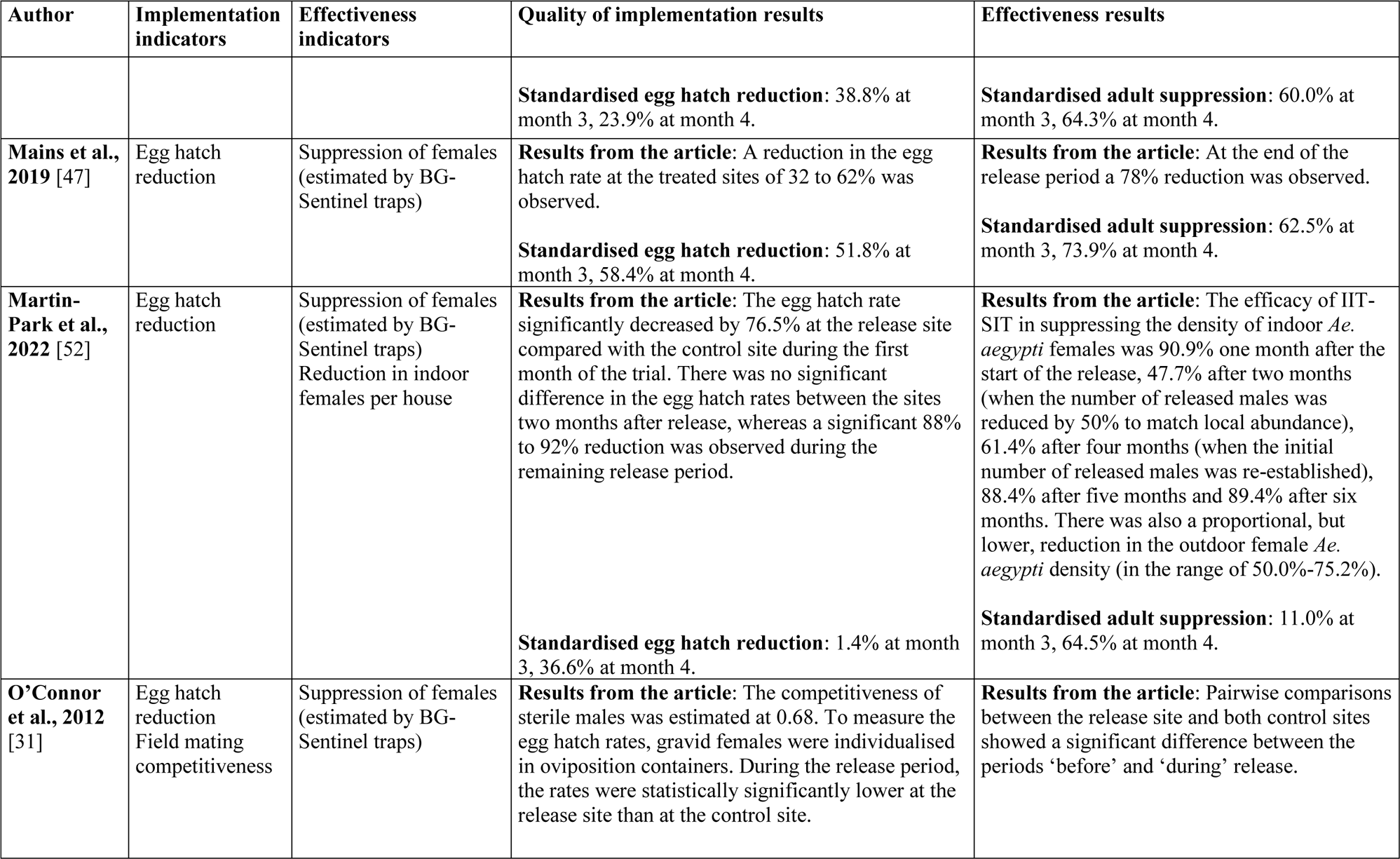
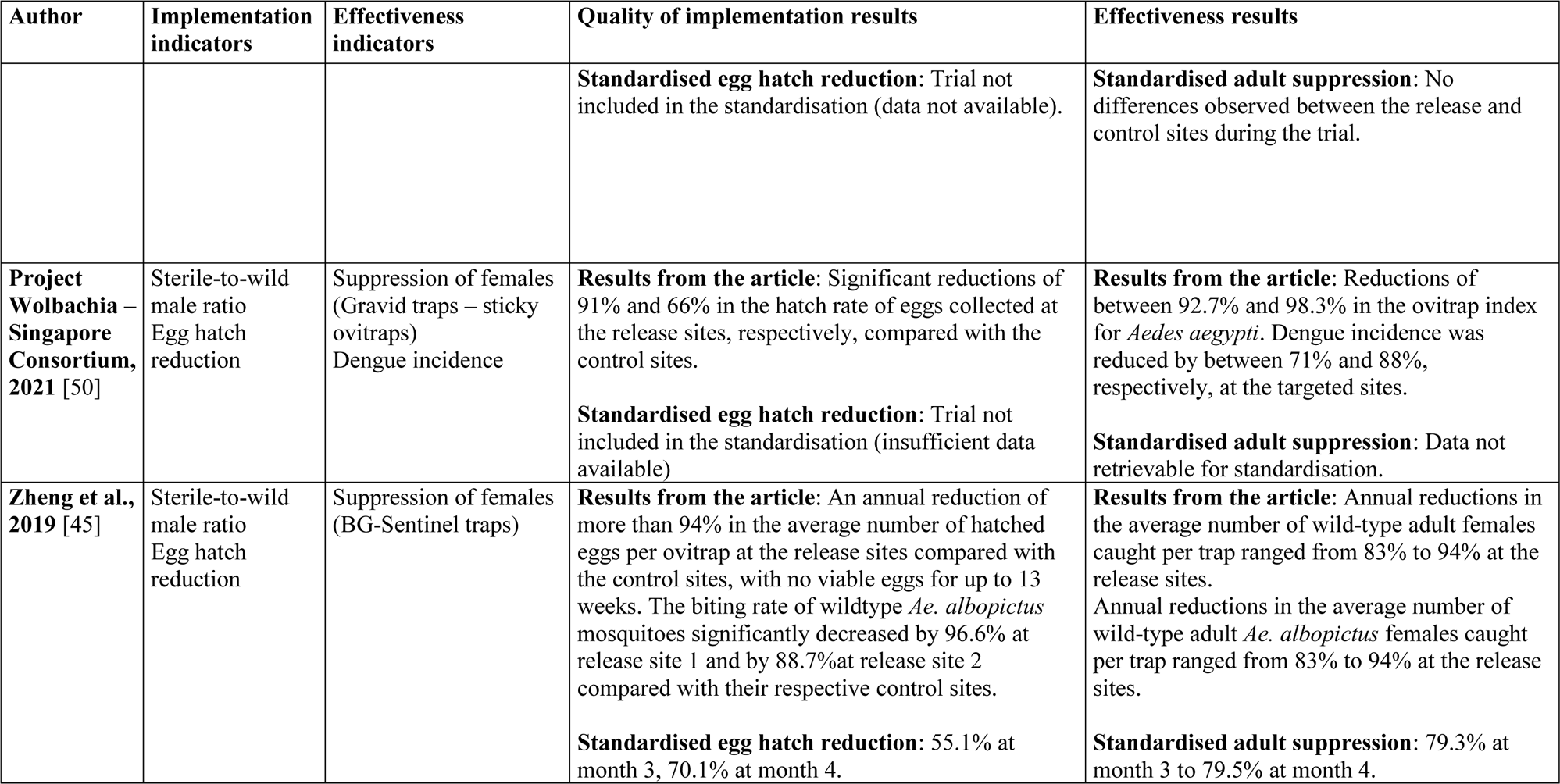
Overview of the main results of the implementation quality (egg hatch reduction) and effectiveness (suppression of females and/or reduction in eggs collected) of the selected SIT, IIT and SIT-IIT trials (n=17).

**Figure 2:**
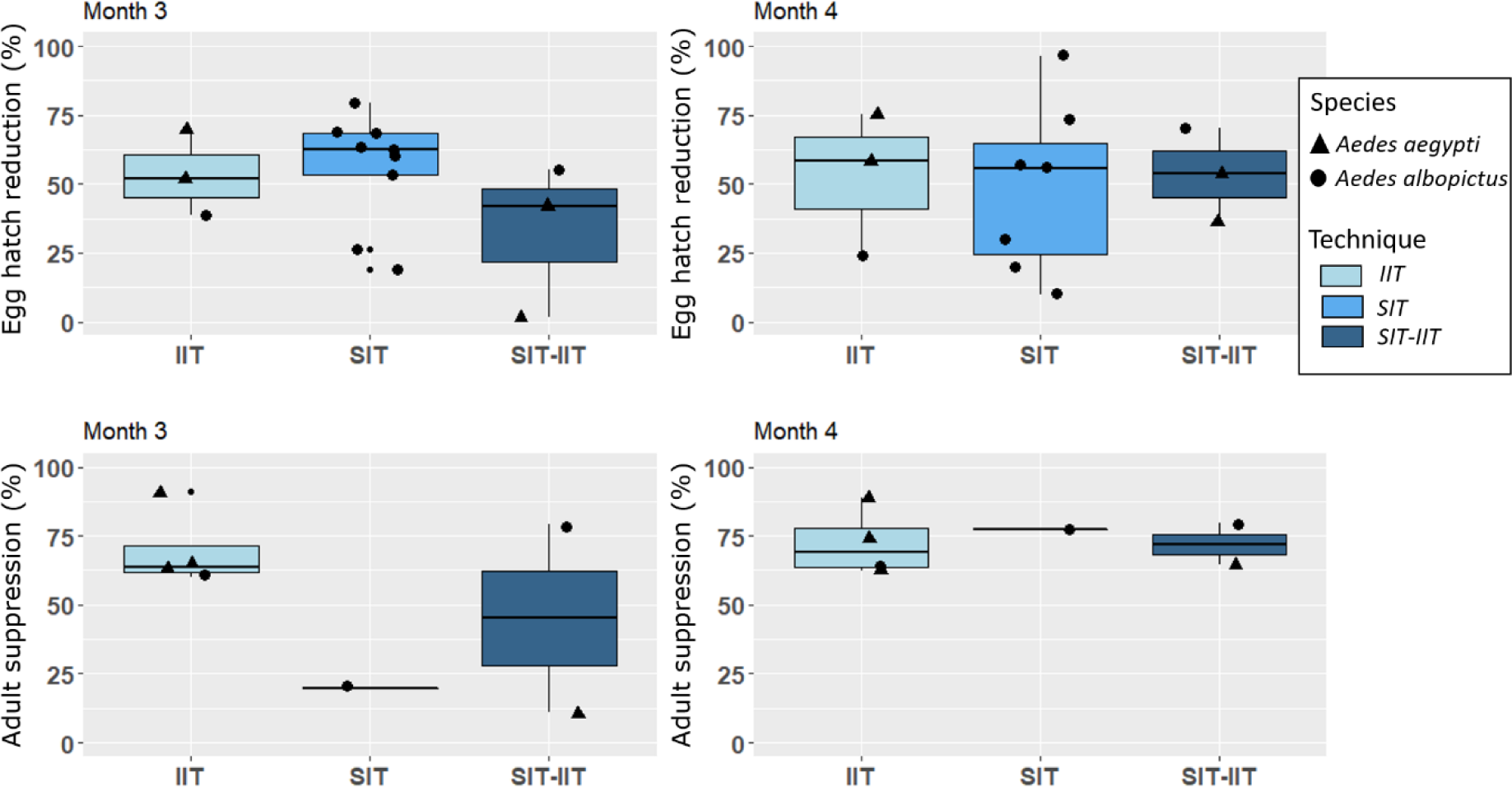
Egg hatch reduction and adult suppression standardised, estimated in the field by ovitraps and BG-Sentinel traps, respectively.

With the limited data available, our analysis suggests a trend in the relationship between egg hatch reduction and female suppression at month 3 after the first release. This is demonstrated by the results of the GLM with a p-value <0.05 and a Pearson correlation coefficient of 0.95 (Figure 3). In addition, the results suggest that when egg hatch reduction is greater than 45% the reduction in females can be as much as 60%, when egg hatch reduction is greater than 60% the reduction females is over 80%, and when egg hatch reduction is greater than 70% the reduction in females is over 90% (Figure 3).

**Figure 3.**
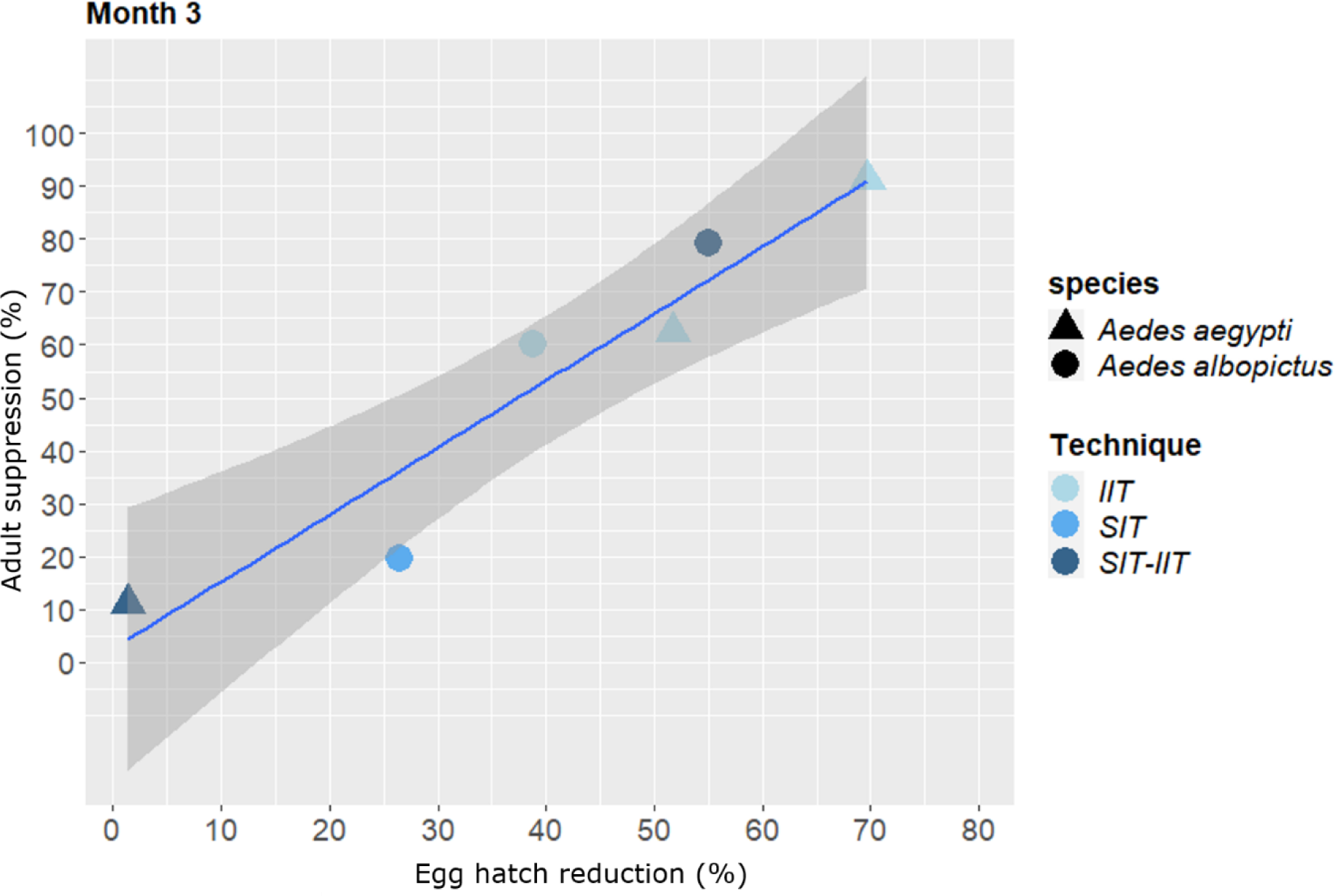
: Relationship between egg hatch reduction and adult female suppression standardised

## Discussion

### Evidence of the effectiveness of the sterile insect and incompatible insect techniques in reducing Aedes female populations

There is evidence that SIT, ITT and SIT-IIT are effective methods for reducing female *Aedes* populations. Indeed, standardised effectiveness indicators show that the reductions are of more than 90% [25,33]. Moreover, our results suggest that effectiveness, i.e. the reduction in adult female mosquitoes, is related to the quality of implementation, i.e. the reduction in the egg hatch rate. More effective implementation will lead to a more effective reduction in *Aedes*, while a substantial egg hatch reduction is needed to radically reduce *Aedes* populations (Figure 2). These results are supported by recent researches that recommend a target of induced sterility of more than 70% to produce additive effects and advise against overcompensation mechanisms that will reduce the effectiveness of these strategies [34]. The results of our standardisation using the available data suggest that the different methodologies (SIT, IIT, SIT-IIT) have comparable levels of efficacy. However, robust studies on the effectiveness of SIT alone are still scarce and they do not allow solid conclusions to be drawn. Confirmation of the effectiveness of this technique may be obtained in the future with the publication of a larger number of field studies.

### Limitations of the study

The first limitation that we acknowledge in this study’s ability to present a global analysis of the effectiveness of the various techniques with solid evidence is underreporting. Indeed, the results of several trials have not yet been published: for example, the IIT trials with *Ae. polynesiensis* in French Polynesia, or the SIT trials with *Ae. albopictus* in Spain [19]. Our analysis is also subject to other limitations, an important one being data availability. The quality of the analysis is weakened by the fact that raw data were not available for the majority of the trials and we had to extract data from the figures (SI table 3). Furthermore, our standardised indicators were based on comparisons between the treatment and control sites and because pre-intervention data were not always available we were unable to use them as the baseline for calculating effectiveness (post-intervention data). We recommend conducting studies in which observations of both the target group and the control group are made before and after implementation of the intervention [35]. Without pre-intervention data, we were of course unable to control for baseline differences introducing potential biases.

### A proposed framework for standardising the effectiveness of SIT, IIT and SIT-IIT in evidence-based vector control trials

To the best of our knowledge, this is the first review of sterile insect and incompatible insect techniques to propose standardisation of indicators, although several meta-analyses have compared the effectiveness of traditional dengue vector control methods [16,36]. A novelty of our study is that, in addition to estimating the effectiveness of three techniques using available information, we also built a framework for standardising evaluation of current evidence presented in published trials. We identified a range of indicators for the sterile and incompatible insect methods, and selected the suppression of adult female abundance as the indicator of effectiveness and the reduction in egg hatch rates as the indicator of implementation quality. Our chosen method for calculating reduction/suppression was Abbott’s formula, which has the advantage of being easy to use with the available data and allowed us to include as many observations as possible in our analysis.

Other indicators merit inclusion in this type of analysis. Although some studies have suggested a relationship between the number of eggs collected and adult suppression [37], others have shown conflicting results [38,39]. Unfortunately, we were unable in our review to address this relationship due to a lack of representative data. Although adult suppression should be a “gold standard”, in some cases egg collection reduction may be a relevant indicator [40]. For example, when there is insufficient data on females catches, egg abundance would be a complementary index to explore. It was not possible to standardise the sterile-to-wild male ratio in our study, nor could we calculate field mating competitiveness, which numerous SIT trials include, since part of the relevant equation is the standard sterile-to-wild male ratio [21,32,41,42].

### Advantages and limitations of SIT, IIT and SIT-IIT

One of the main advantages of sterile and incompatible insect control methods is that they are species-specific and non-insecticidal. They are, therefore, environmentally friendly and have a limited impact on biodiversity and non-target insect species, such as pollinators and biological enemies. In addition, the SIT technique has already been successfully used to control agricultural pests, such as the New World screwworm, which has been eliminated in North and Central America, and fruit flies [43]. SIT and IIT are most cost-effective when used as part of an integrated vector management programme, as suggested by trials in Germany and Mauritius [21,44,45]. Although the importance of community engagement was not explored in this review, community participation, acceptability and intersectoral coordination are crucial to the success of these techniques [34]. This is particularly true for the more traditional *Aedes* control techniques: without community acceptability and support there is a greater risk of ineffective implementation and failure. For the SIT and IIT techniques, acceptance and engagement appear to be very important in the context of integrated vector management, particularly during maintenance phases when the level of release is lower and monitoring and prevention of reinvasion are essential [46]. Furthermore, female contamination, that is, the migration of mosquitoes from surrounding (untreated) sites would limit the sustainability and effectiveness of this method in the long term [18]. We consider data from experimental studies evaluating synergies with other vector control methods, such as community engagement, source reduction strategies, larvicide and mass trapping, to be essential in improving the long-term effectiveness of these methods. Importantly, unlike some control strategies, such as indoor/outdoor insecticide spraying or door-to-door source reduction, the SIT and IIT techniques do not require access to private property, but are, instead, complementary to strategies requiring community participation [44]. This is an important advantage, as an optimal implementation will provide optimal control coverage and therefore a potentially effective intervention. It is important to bear in mind that public acceptability is a mandatory requirement for the implementation of these strategies [20].

A major challenge for the release of sterile and incompatible males is the quality of the males themselves and, in particular, their sexual competitiveness [32]. The lower quality of the sterile male insects produced could be related to several factors, such as irradiation, but also the processes of mass rearing, handling, transport and release. All these processes require significant infrastructure and expertise as well as financial investment, although they also create employment and stimulate innovation and development. There is a lack of data on the economic costs of these interventions, making it impossible to calculate their cost-effectiveness.

SIT and IIT are suitable strategies for routine vector control, but not as emergency measures as they take several months to be fully effective, with implementation ideally beginning early in the mosquito season [34]. In an epidemic emergency, controlling an arbovirus would require rapid interruption of the transmission cycle (within weeks) and, so far, only insecticidal methods are capable of this. Moreover, legislative issues are a major consideration. Although IIT or SIT-IIT have been shown to be effective in trials in the USA, China, Australia, Singapore and Thailand, they cannot be applied in all settings due to local legislation.

Regarding the use of *Wolbachia* to reduce arbovirus transmission, it is important to point out that our review only covered population suppression strategies. There are also “population replacement” strategies that use the *Wolbachia* method (e.g. the World Mosquito Program; https://www.worldmosquitoprogram.org/). As the unintentional release and establishment of *Wolbachia* in target populations poses an epidemiological risk that would compromise IIT control methods [46], an important part of these methods should be to monitor for such an event [25,28,47].

### Recommendations for standardisation of future trials

We recognise that evaluating egg hatch reduction, adult suppression and reduced egg collection require significant human, logistical and economic resources. However, we recommend that all these indicators be evaluated, where possible, to provide robust evidence of the effectiveness of these methods. In general, studies evaluating the effectiveness of vector control interventions lack rigour in key aspects such as design, implementation, and data management and analysis [16]. We have offered guidance to improve the overall operation of field trials of vector control tools and strategies [48]. To be able to compare different trials, the data should be made available in peer-reviewed scientific publications, but also, where possible, in a common, standardised database. This is currently the case with some studies, such as the trials in California and Thailand [25,33]. A living database of *Aedes* control trials using, for example, the sterile and incompatible insect techniques, but also other innovative methods (e.g. mass trapping, pyriproxyfen autodissemination) with one or more standardised indicators would be helpful for comparing different methods. This would support further proof-of-concept of innovative and traditional *Aedes* control methods, that are increasingly important but challenging. The second requisite is to adopt several common indicators in all trials, which could be used to generate information on the quality of the technique’s implementation and evidence of its effectiveness. In this study, we proposed using egg hatch rates for implementation quality and female adult abundance for effectiveness. Calculating these indicators requires the data collection method (e.g. trapping) to be tailored. Other indicators would also be relevant for comparison purposes, such as the ratio of sterile to wild males and the mating competitiveness of sterile males. Having these indicators in common would allow other issues to be explored, such as the relationship between male competitiveness and the effectiveness of a method. It is important for any field trial evaluating the effectiveness of a vector control method to have a robust experimental design. Contemporary comparison sites (i.e. untreated), pre-treatment (baseline) measures, randomised selection of treated/untreated sites, calculation of a sufficient number of clusters and appropriately sized sites calculated with power analysis are essential elements [35]. In some cases, using a buffer zone and a “fried egg” design can reduce potential biases due to contamination by females from outside the treated sites [49]. In assessing the potential cost-effectiveness or cost-benefits it is important to report the economic costs of the intervention. Finally, until now the sterile and incompatible insect techniques have been tested on a small scale in pilot studies. With the exception of the Singapore trial [50], few studies have investigated the impact of these techniques on epidemiological outcomes and arbovirus transmission. It remains to be demonstrated that these techniques can be scaled up as effective operational tools for disease control and the reduction of arbovirus transmission [18].

## Conclusion

This review has proposed standardised indicators − egg hatch reduction and female suppression − for field trials in order to facilitate assessment and comparison of the effectiveness of SIT, IIT and SIT/IIT against *Aedes* mosquitoes. Our results suggest that, when implemented effectively, the incompatible and sterile insect techniques are highly effective in suppressing *Aedes* mosquito populations. Our analysis is limited by data availability and underreporting of trial results. In addition to their potential effectiveness, these techniques are species-specific, non-insecticidal and environmentally friendly. However, it has not yet been shown that the sterile insect technique and the incompatible insect technique can be sufficiently scaled up to be effective operational tools for reducing arbovirus transmission.

## Data Availability

All data produced in the present study are available upon reasonable request to the authors

## Acknowledgements and funding

This review is part of the project “Convention de Recherche et Développement” No. 2019-CRD-12 funded by ANSES (Agence Nationale de Securité Sanitaire de l’Alimentation, de l’Environnement et du Travail).

## Availability of data and materials

All the data standardized are provided in the manuscript (Table 2).

## Authors’ contributions

MMO: conceived and designed the review, screened titles and abstracts, extracted data, standardised indicators and wrote of the first version of manuscript

GLG: conceived and designed the review, screened titles and abstracts, validated data extraction and wrote the manuscript

TB: conceived and designed the review and wrote the manuscript

DR: conceived and designed the review, wrote the manuscript and supervised of the study

