## Supplemental tables and figures for "Are the sterile insect technique and the incompatible insect techniques effective in reducing *Aedes* mosquito populations?"

SI Table 1. Inclusion and exclusion criteria

| Item | Inclusion criteria | Exclusion criteria |
| --- | --- | --- |
| Species | *Aedes aegypti* OR *Aedes albopictus* OR *Aedes polynesiensis* | Other mosquito species |
| Vector control techniques | SIT  IIT  SIT-IIT  Boosted SIT | Wolbachia Aedes species replacement strategies.  Other vector control methods, such as insecticide, larvicide, traps, RIDL, transgenesis |
| Study design | Field study  Controlled study | Laboratory study  Male production study  Release-recapture study  Semi-field study  Mathematical modelling  Theorical study  Opinion article  Review |
| Target | Adult  Egg | Pupae  Larvae  Adult humans |
| Index description /outcomes | Adult density  Egg hatch rate  Egg density | Mosquito biting rate  Larval indices (e.g. house index, container index, Breteau index, pupae indices)  Vector infection  Qualitative study |

**SI Table 2:** Overview of the experimental design of the 17 SIT, IIT and SIT-IIT trials

| **Author** | **Type of publication** | **Study site location, country** | **Year of trial** | **Technique** | **Targeted species** | **Epidemiological context** | **Extent of the intervention area** | **Average number of sterile males released per week per ha standardised (total released; source of data)** | **Duration of the release** |
| --- | --- | --- | --- | --- | --- | --- | --- | --- | --- |
| **Balastos et al., 2021** | Peer-reviewed article | Vravrona and Markopoulo municipalities, Attica, Greece, | 2018 | SIT | *Aedes albopictus* | Nuisance and imported cases of dengue. Historic dengue outbreak (1927–1928). | 5 ha | 2,649 (18,542; publication) | 10 weeks |
| **Becker et al., 2022** | Peer-reviewed article | Melm district Ludwigshafen city, Germany  Metzgergrun, Freiburg city, Germany | 2020 | SIT | *Aedes albopictus* | Ongoing invasion of the Asian tiger mosquito in the Rhine valley.  Imported cases of dengue. | Melm 17 ha + Metzgergrün 4.5 ha | Melm area: 1,013 (310,000; publication) Metzgergrün area: 2,320 (136,000; publication) | Melm: 18 weeks; Metzgergrün: 15 weeks |
| **Bebee et al., 2021** | Peer-reviewed article | 3 replicates Cassowary Coast Region, Queensland, Australia | 2018 | IIT | *Aedes aegypti* | Nuisance and imported cases and autochthonous transmission of dengue (Akter et al., 2019) | 3 replicates: 44, 65 and 85.5 ha, respectively | 2,170 (2,800,000; estimated from publication) | 20 weeks |
| **Bellini et al., 2013** | Peer-reviewed article | Santamonica, Italy  Boschi, Italy  Budrio, Italy  Caselline, Italy | Santamonica: 2005  Boschi: 2008 and 2009  Budrio: 2008  Caselline: 2009 | SIT | *Aedes albopictus* | Nuisance and imported cases of dengue.  Outbreaks of dengue in 2020 (Lazzarini et al., 2020) and chikungunya in 2007 (Angelini, 2007) and 2017 (Rezza, 2018) | Santamonica: 45 ha  Boschi: 16 ha  Budrio: 17 ha  Caselline: 18 ha | Santamonica: 830 (525,000; publication)  Boschi, 2008: 1,680 (600,000; publication)  Boschi, 2009: 1,140 (310,000; publication)  Budrio: 950 (360,000; publication)  Caselline: 628 (341,000; publication) | Santamonica: 14 weeks  Boschi: 22.3 weeks (2008) and 17 weeks (2009)  Budrio: 14.3 weeks  Caselline: 30 weeks |
| **Bellini et al., 2021** | Peer-reviewed article | Boschi, Italy  Budrio, Italy  Caselline, Italy  Gherghenzano, Italy  Caselle, Italy  Guisa Pepoli, Italy | Santamonica: 2005  Boschi: 2008 and 2009 (duplicate Bellini and al., 2013) and 2010  Budrio: 2008 (duplicate Bellini and al., 2013)  Caselline: 2009 (duplicate Bellini and al., 2013)  Gherghenzano: 2012  Caselle: 2018  Guisa Pepoli: 2018 | SIT | *Aedes albopictus* | Nuisance and imported cases of dengue.  Outbreaks of dengue in 2020 (Lazzarini et al. 2020) and chikungunya in 2007 (Angelini, 2007) and 2017 (Rezza, 2018) | Gherghenzano: 10 ha  Caselle: 16 ha  Guisa Pepoli: 7 ha | Boschi, 2010: 860 (240,000; estimation)  Gherghenzano, 2012: 10,700 (2,025,000; estimation)  Caselle, 2018: 226 (118,000; estimation)  Guisa Pepoli, 2018: 780 (179,000; estimation) | Boschi 2010: 18 weeks  Gherghenzano: 19 weeks  Caselle: 33 weeks  Guisa Pepoli: 33 weeks |
| **Bouyer et al., 2020** | Peer-reviewed article | Carnaiba do Sertao, Juazeiro, Brazil | 2018 | SIT | *Aedes aegypti* | Endemic circulation of dengue.  Outbreaks of chikungunya (2014, 2015) and Zika (2016-2017) | 20 ha | Phase 2 (drone release along release lines 80 m apart): 5,000 (165,400; publication) | 2 or 3 releases over one week then a 3-week observation period |
| **Caputo et al., 2019** | Peer-reviewed article | Urban Rome, Italy | 2018 | IIT | *Aedes albopictus* | Nuisance and imported cases of dengue.  Outbreaks of dengue in 2020 (Lazzarini et al., 2020) and chikungunya in 2007 and 2017 (Rezza, 2018) | 2.7 ha | 1,700 (26,680; publication) | 6 weeks |
| **Crawford et al., 2020** | Peer-reviewed article | 3 replicates:  Fresno County, California, USA | 2018 | IIT | *Aedes aegypti* | Nuisance, imported and autochthonous cases of dengue, Zika and chikungunya | 3 replicates:  293 ha | 3 replicates with an average of 1,885 males per hectare per week (14.4 million males across three replicates; publication | 26 weeks |
| **Gato et al., 2020** | Peer-reviewed article | Havana city, Cuba | 2020 | SIT | *Aedes aegypti* | Endemic circulation of dengue | 50.2 ha | 1,200 (1,270,000; publication) | 21 weeks |
| **Iyaloo et al., 2019** | Preprint | Panchvati and Pointe des Lascars,  Mauritius | 2017-2018 | SIT | *Aedes albopictus* | Recurrent dengue outbreaks (2009, 2014, 2015, 2019 and 2020). Chikungunya outbreak (2005-2006) | 3 ha | 19,700 (2,250,000; publication | ~ 38 weeks |
| **Kittayapong et al., 2019** | Peer-reviewed article | Plaeng Yao district, Chachoengsao province, Thailand | 2016 | SIT-IIT | *Aedes aegypti* | Endemic circulation of dengue | 65 ha | 280 (437,980;) | 24 weeks |
| **Mains et al., 2016** | Peer-reviewed article | Lexington, Kentucky, USA | 2014 | IIT | *Aedes albopictus* | Nuisance and imported cases of dengue.  Recurrent dengue outbreaks (Florida, Texas, Hawaii).  Outbreaks of Chikungunya (2014, 2015) and Zika (2016-2017) | The treated site is reported as having a 250 m radius around a single release point (treatment area estimated at ~12.5ha) | 850 (182,000; publication) | 17 weeks |
| **Mains et al., 2019** | Peer-reviewed article | Miami,  USA | 2018 | IIT | *Aedes aegypti* | Nuisance and imported cases of dengue.  Recurrent dengue outbreaks (Florida, Texas, Hawaii).  Outbreaks of chikungunya (2014, 2015) and Zika (2016-2017) | ~170 acres ~68,8 ha | 5,450 (6.8 million; publication) | 24 weeks |
| **Martin-Park et al., 2022** | Peer-reviewed article | Yucatan,  Mexico | 2019 | SIT-IIT | *Aedes aegypti* | Endemic circulation of dengue.  Outbreaks of chikungunya (2014, 2015) and Zika (2016-2017) | 50 ha | 2,100 (1,270,000; publication) | 24 weeks |
| **O'Connor et al., 2012** | Peer-reviewed article | Tiano, Horea and Toamari islands,  French Polynesia | 2009-2010 | IIT | *Aedes polynesiensis* | Recurrent dengue outbreaks.  Outbreaks of chikungunya (2014, 2015) and Zika (2013-2014).  Lymphatic filariasis | Not specified in the study | 3,800 males per week (117,000;) | 30 weeks |
| **Project Wolbachia – Singapore Consortium, 2021** | Preprint | Singapore | Phase 2: 2019 | Phase 1: IIT  Phase 2 (suppression trial): IIT in Yishun; SIT-IIT in Tampines (but using a high-fidelity sex-sorting pipeline) | *Aedes aegypti* | Endemic circulation of dengue | Not clearly specified; Yishun around 4 ha, Tampines around 2 ha | The data available in the preprint paper do not allow a standardised indicator to be calculated | Phase 1: 15 and 31 weeks Phase 2: 17 months |
| **Zheng et al., 2019** | Peer-reviewed article | Guangzhou, China | 2016-2017 | SIT-IIT | *Aedes albopictus* | Endemic circulation of dengue | Site 1: 25 ha Site 2: 7.5 ha | 90,600 (5,590,000; publication) | 20 months |

SI Table 3. Reliability of the indicators standardised

| Reference | Country | Quality of the data extracted |
| --- | --- | --- |
| Balastos et al., 2021 | Greece | Low |
| Becker et al., 2022 | Germany | Moderate |
| Beebe et al., 2021 | Australia | Low |
| Crawford et al., 2020 | USA | High |
| Kittayapong et al., 2019 | Thailand | High |
| Gato et al., 2020 | Cuba | Low |
| Iyaloo et al., 2019 | Mauritius | Low |
| Main et al., 2016 | USA | Low |
| Main et al., 2019 | USA | Low |
| Martin-Park et al., 2022 | Mexico | Low |
| Zheng et al., 2019 | China | Low |
