## Supplementary figures and images for "Are the sterile insect technique and the incompatible insect techniques effective in reducing *Aedes* mosquito populations?"

### Supplemental figure 1

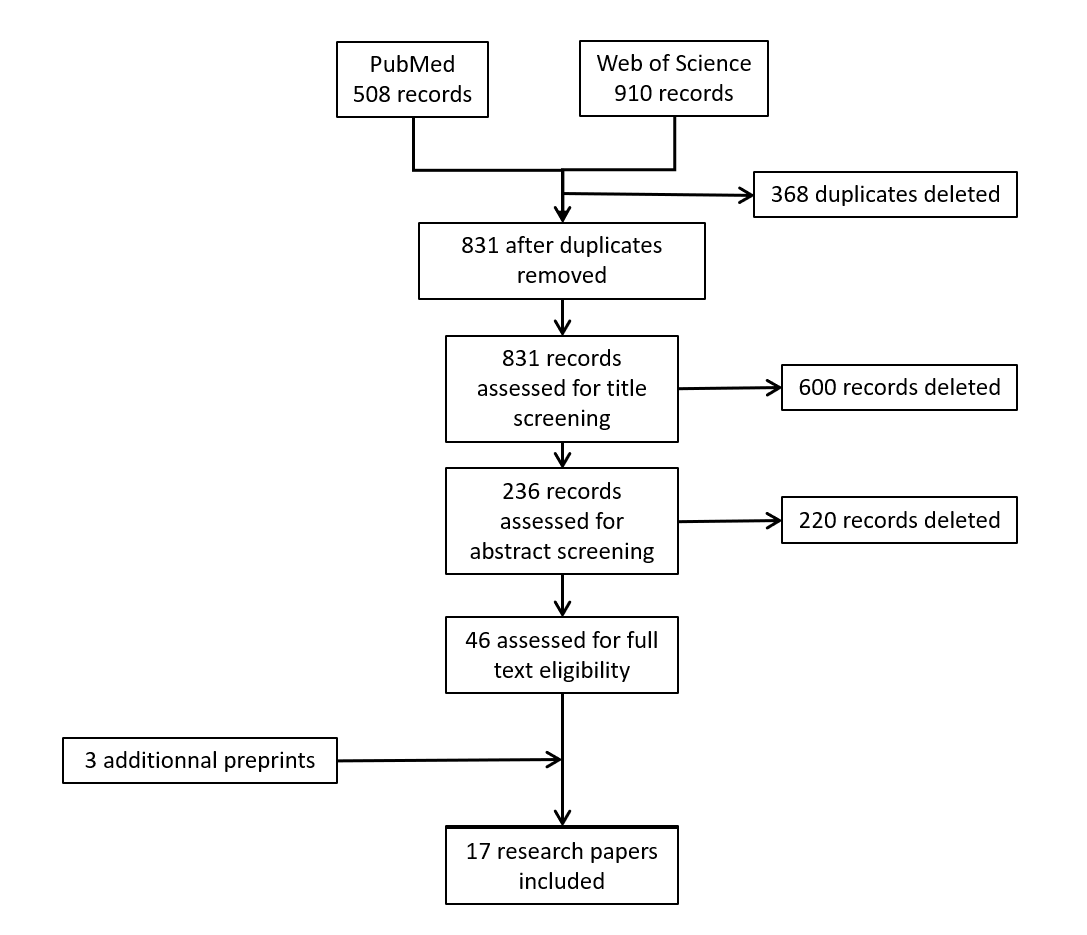
